# Reconstructed diagnostic sensitivity and specificity of the RT-PCR test for COVID-19

**DOI:** 10.1101/2020.04.24.20078949

**Authors:** N. S. Padhye

## Abstract

Real-time reverse transcription polymerase chain reaction (RT-PCR) targeting select genes of the SARS-CoV-2 RNA has been the main diagnostic tool in the global response to the COVID-19 pandemic. This study was aimed at the estimation of diagnostic sensitivity and specificity of the first RT-PCR test developed by China CDC in January 2020. The study design is a secondary analysis of published findings on 1014 patients in Wuhan, China, of whom 59.3% tested positive for COVID-19 in RT-PCR tests and 87.6% tested positive in chest CT exams. We utilized previously ignored expert opinions in the form of verbal probability classifications of patients with conflicting test results to estimate the informative prior distribution of the infected proportion. It was then used in a Bayesian version of a previously developed model to reconstruct the sensitivity and specificity of the diagnostic tests without the need for specifying an inaccurate test as the gold standard. The sensitivity of the RT-PCR diagnostic test was estimated to be 0.707 (95% CI: 0.668, 0.749), while the specificity was 0.851 (95% CI: 0.774, 0.941). Caution is advised in generalizing these findings to other versions of the RT-PCR test that are being used in diverse geographic regions.

## Introduction

The cause of a disease outbreak that began in Wuhan, China in the last quarter of year 2019 was later identified as a novel coronavirus, labeled SARS-CoV-2 since it can cause severe acute respiratory syndrome, a disease that has been named COVID-19 [33]. The publication of the SARS-CoV-2 genome [30] led to the rapid development in January 2020 of real-time reverse transcription polymerase chain reaction (RT-PCR) tests for the diagnosis of COVID-19 while avoiding cross-reactions to other known coronaviruses. Among early versions, one was developed in China that targeted the *ORF1ab* and *N* genes of viral RNA [34] while another version was developed in Germany that targeted the *RdRp, E*, and *N* genes [8]. Real-time RT-PCR tests were developed and implemented thereafter by many laboratories around the world [6, 19, 24], even as COVID-19 became a global pandemic that continued to spread rapidly. A listing of tests and protocols is maintained online by the World Health Organization [35].

The rapid development and deployment of RT-PCR tests has been essential for the ability to measure and control the spread of SARS-CoV-2. However, the urgency of the pandemic has meant that the diagnostic tests were deployed without first being supported by clinical studies to measure the diagnostic error rates. Initial knowledge about the RT-PCR tests was based on laboratory measurements of sensitivity to the minimum threshold of detection of viral loads and the required number of thermal cycles of the chain reaction [21]. Soon after deployment of the tests, attention was given to the viral distribution by physical location, such as the differences in positive rates of RT-PCR in nasopharyngeal versus oropharyngeal swabs, or in the sputum and bronchoalveolar lavage fluid [27, 31]. Other factors that can impact the diagnostic success of RT-PCR include the timing of the test relative to disease onset, adequacy of the volume of fluids collected in the swab, and deviations from the laboratory-recommended protocol under real-world conditions. In terms of clinical decision-making, any of the causes of failure of the test led to incorrect diagnoses that resulted in false positives or false negatives and the lack of reliable data on diagnostic accuracy received media attention [12, 15] as the pandemic unfolded.

This study provides an assessment of the diagnostic sensitivity and specificity of the RT-PCR diagnostic test that was used shortly after the pandemic began in Wuhan, China. The data were collected in a study that had the aim of measuring the accuracy of chest computerized tomography (CT) imaging for diagnosis of COVID-19 in 1014 patients [1] and the investigators assumed that RT-PCR was the gold standard. The study authors provided additional information about the status of patients in the form of verbal probabilities of infection that were assigned after a review. Thus, in addition to the counts of positives and negatives for chest CT and RT-PCR, there were unutilized data in the form of expert judgments about the patients. In this study, it is shown that putting together all of the available data permits the reconstruction of the sensitivity and specificity of RT-PCR for clinical decision-making without the need for a gold standard test. A by-product of this approach resulted in the assessment of diagnostic performance of chest CT imaging.

There are two main features of the methods used in this study. The first feature is the use of fuzzy membership functions that creates a pathway for the inclusion of verbal probability data that would otherwise be ignored. The second feature is the use of the Bayesian form of modeling developed originally by Dawid and Skene in the maximum likelihood framework [9] that does not need the existence of a gold standard test for evaluation of the accuracy of a diagnostic test. However, it does need an informative prior distribution for the proportion of infected cases, which is possible to estimate from the verbal probability data. The two features are therefore related with the second feature arising as a result of the first one. The ability to discount the need for a gold standard test is important because neither the RT-PCR test nor the chest CT test are accurate enough to be considered gold standard tests for COVID-19. Reverse calculations that rely on considering one or another test as gold standard produce biased estimates, which can be avoided in the approach adopted here.

## Methodology

### Data

The methods were applied to data from a study that retrospectively enrolled patients suspected of having COVID-19 who underwent RT-PCR and chest CT imaging diagnostic tests at Tongji Hospital of Tongji Medical College of Huazhong University of Science and Technology in Wuhan, Hubei, China, during a 30-day period in the months of January and February, 2020 [1]. The effective sample size was 1014 and it was reported that 46% were male while the mean age was 51 ±15 years. Throat swab samples were collected and the RT-PCR assays were reported to have used TaqMan One-Step RT-PCR kits from Shanghai Huirui Biotechnology Co., Ltd., or Shanghai BioGerm Medical Biotechnology Co., Ltd., both of which were approved for use by China Food and Drug Administration. Chest imaging was done on one of three CT systems at the hospital and two radiologists reviewed the images while being blinded to the molecular test results. The median time interval between the chest CT exams and RT-PCR assays was 1 day.

RT-PCR assays tested positive for 601 patients (59.3%) and negative for the other 413 patients (40.7%). Chest CT exams were positive for 888 patients (87.6%) and negative for the other 126 patients (12.4%). See Table 1 for the joint distribution of the two tests. A large block of 308 patients with conflicting test results were reassessed on the basis of clinical symptoms and serial CT scans. The investigators concluded that 147 of these patients could be classified as *highly likely* cases of COVID-19 and another 103 could be classified as *probable* cases of COVID-19. Patients in both classifications had clinical symptoms of COVID-19, but repeat CT scans showed progression of disease in the *highly likely* cases while being stable in the *probable* cases. In summary, the data include the joint distribution of test results from RT-PCR and chest CT along with expert opinion in the form of verbal probabilities.

**Table 1.** Summary of data for the observed joint distribution of RT-PCR and chest CT test results. See the footnote for a summary of clinical expert opinion data.

|  | Positive chest CT | Negative chest CT | Row Sums |
| --- | --- | --- | --- |
| Positive RT-PCR | 580 | 21 | 601 |
| Negative RT-PCR | 308* | 105 | 413 |
| Column Sums | 888 | 126 | 1014 |

\*Expert opinion indicated that COVID-19 was *highly likely* in 147 of these cases and *probable* in another 103 cases.

### Statistical Analysis

Dawid and Skene developed a maximum likelihood model for data from multiple raters that rigorously accounted for the uncertainty of the true rating of a case [9]. Their model is used here in its Bayesian form for estimation of the sensitivity and specificity of diagnostic tests. The RT-PCR and chest CT diagnostic tests are analogous to two raters that provide binary ratings and the true diagnosis, *z*_*i*_ = 0 or 1 for the *i*^th^ patient is unknown in the absence of a gold standard test for COVID-19. In order to estimate the informative prior distribution of the proportion of patients with COVID-19, it was necessary to use ideas from fuzzy logic that are described below in addition to details about the implementation of the Bayesian model.

Linguistic uncertainties associated with terms such as *highly likely* and *probable* can be represented by membership functions of fuzzy logic that are not to be confused with probability distributions. A membership function can be defuzzified to yield a crisp estimate [32]. In the current application, the defuzzification yielded a probability associated with the linguistic terms. A recent study provided detailed estimates of membership functions of commonly used verbal probabilities [28]. In order to use their membership functions it was necessary to make a correspondence between the terminology used in the two studies [1, 28]. Equivalence was assumed between the terms *highly likely* and *very likely*, while the term *probable* was considered equivalent to *likely*, which is supported by synonyms for *probable* that are listed in the thesaurus by Oxford Languages. The centroids of membership functions yielded the values of the probability of being a *highly likely* or a *probable* case of COVID-19. Thus, the defuzzification provided a pathway to an exclusively probabilistic approach in the subsequent computation without having altogether ignored the linguistic uncertainties.

The estimated verbal probabilities *p*_*v*_ parametrized the Bernoulli distribution B(*p*_*v*_) that was used for imputation of ten thousand samples, each of size equal to the original data. Note that values of *p*_*v*_ for a patient could be 0 or 1 in addition to the verbal probabilities that were estimated for the *highly likely* and *probable* classifications. The imputation resulted in the estimated distribution *π* of the proportion of patients that presented with COVID-19. For convenience of the Bayesian implementation in *RStan* [26], the informative prior for *π* was approximated by a beta distribution after matching the 2.5% and 97.5% quantiles to the imputed distribution using the *Beta*.*Parms*.*from*.*Quantiles* program [3] in *R*.

The remaining parameters of the model may be denoted by *θ*_*j,k*_ for the probability of correct or incorrect assignment by the *j*^th^ type of diagnostic test for the *i*^th^ patient with true diagnosis *z*_*i*_ = *k*, where *k* is binary. The parameters of primary interest in this study are *θ*_*j,k*_ since they represent sensitivity and specificity for each type of diagnostic test. The true diagnosis, *z*_*i*_ = *k*, was marginalized over *k* in the *RStan* implementation because of its inability to sample discrete parameters. Thus, the Hamiltonian sampling was designed to estimate the logarithmic form of

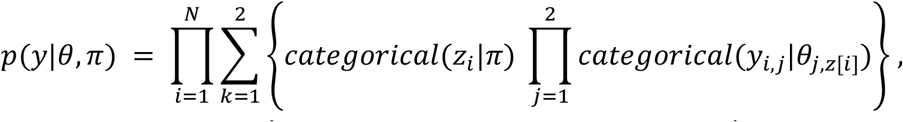

where *y*_*i,j*_ denotes the observed *j*^th^ type of diagnostic test result for the *i*^th^ patient. Although the discrete parameter was marginalized, estimates were obtained of the probabilities of the true diagnosis, *p*(*z*_*i*_|*θ, π*). Noninformative prior distributions were used for the sensitivity of both tests: beta distributions with shape parameters set equal to half, i.e. *Beta*(0.5, 0.5). Weakly informative prior distributions were used for the specificity of tests to assist with model convergence and to avoid inappropriate inferences. The 2.5% and 97.5% quantiles of the beta distributions for specificities covered a wide span between 0.50 and 0.95. Four chains were used while running the model in *RStan*, with 2500 iterations per chain that were run in parallel on four cores of an Intel i5 processor.

Lastly, the positive and negative predictive values of each test were evaluated. These provide the chance of disease in a patient conditional upon results of the diagnostic test, i.e. P(COVID-19 | Positive test result) or P(COVID-19 | Negative test result). It is straightforward to relate the predictive values to the sensitivity, specificity, and pre-test probabilities of infection using Bayes’ rule. Statistical analysis was done using the *R* programming language [23] in the *RStudio* software environment [25].

## Results

Centroid defuzzification resulted in probability values 0.843 and 0.746 for *highly likely* and *probable* cases of COVID-19, respectively. Imputation of 10,000 samples based on Bernoulli trials indicated that the prior distribution of the proportion of patients who presented with COVID-19 had a median of 0.791 while the quantiles corresponding to 2.5% and 97.5% were 0.778 and 0.803. The beta distribution with matched quantiles had shape parameters *a* = 876.2 and *b* = 3307.4 and its mode matched within a margin of 0.00025, thereby providing a good substitute that acted as the informative prior for the Bayesian calculation in *RStan*.

The convergence and mixing of chains were satisfactory, resulting in estimates of 0.707 for the sensitivity and 0.851 for the specificity of the RT-PCR test. In contrast, the sensitivity for chest CT was estimated to be quite high at 0.992, but it had a low specificity value of 0.595. See Table 2 for error estimates and more details. The model also provided estimates of the probability of having COVID-19 for each patient and these are shown in Table 3 for each of four combinations of RT-PCR and chest CT test results. The values ranged from a low of 0.016 when both tests were negative to a high of 0.978 for two positive tests. In the scenario of conflicting test results, the probability of infection was higher for a positive chest CT exam than for a positive RT-PCR test.

**Table 2.**
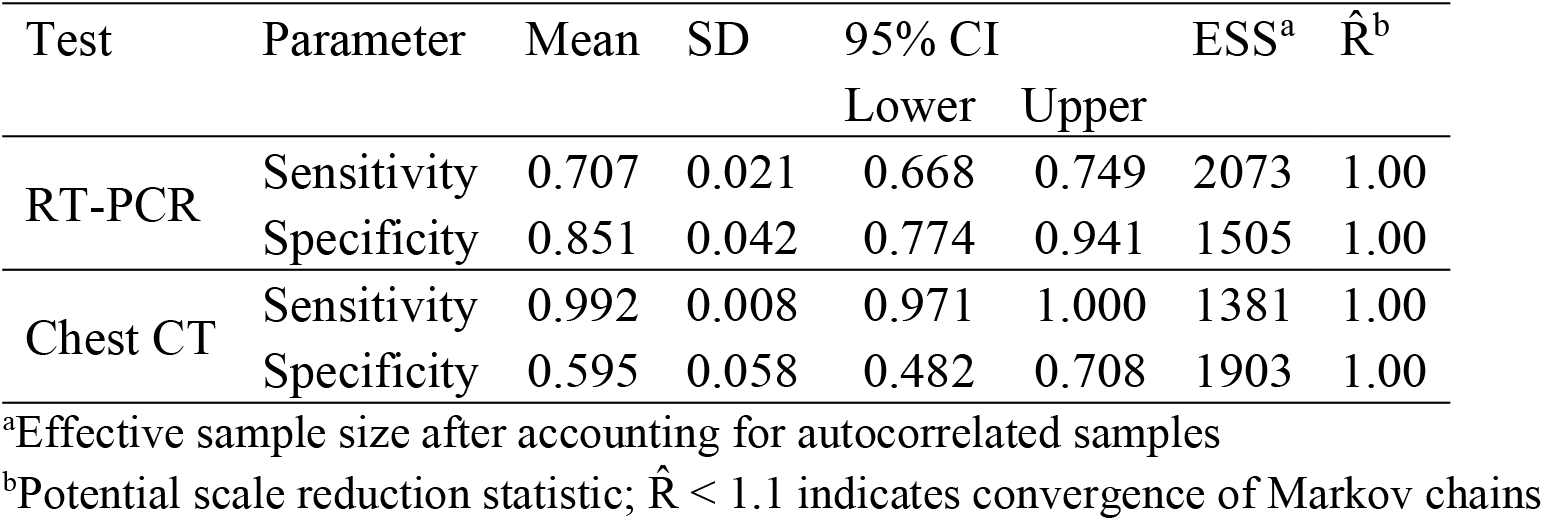
Estimated sensitivity and specificity of RT-PCR and chest CT diagnostic tests for COVID-19.

**Table 3.** Estimated probabilities of COVID-19 for combinations of RT-PCR and chest CT diagnostic test results.

| RT-PCR | Chest CT | Mean | SD | 95% CI | | ESS<sup>a</sup> | $\hat{R}^b$ |
| --- | --- | --- | --- | --- | --- | --- | --- |
|  |  | Probability |  | Lower | Upper |  |  |
| Negative | Negative | 0.016 | 0.017 | 0.000 | 0.062 | 1453 | 1.00 |
|  | Positive | 0.761 | 0.041 | 0.670 | 0.834 | 1442 | 1.00 |
| Positive | Negative | 0.187 | 0.197 | 0.000 | 0.693 | 1085 | 1.01 |
|  | Positive | 0.978 | 0.006 | 0.965 | 0.991 | 1868 | 1.00 |

<sup>a</sup>Effective sample size after accounting for autocorrelated samples
<sup>b</sup>Potential scale reduction statistic; $\hat{R} < 1.1$ indicates convergence of Markov chains

The predictive values of the RT-PCR and chest CT diagnostic tests are shown in Figure 1 for prior (pre-test) probabilities ranging from 0 to 1. The two curves in each panel show the posterior (post-test) probabilities of the presence of COVID-19 when test results are either positive or negative. It can be seen that the post-test probability of infection is generally higher for a positive RT-PCR test than for a positive chest CT, which is related to the higher specificity of RT-PCR. On the other hand, the post-test probability of infection is lower when chest CT is negative because of its higher sensitivity.

**Figure 1.**
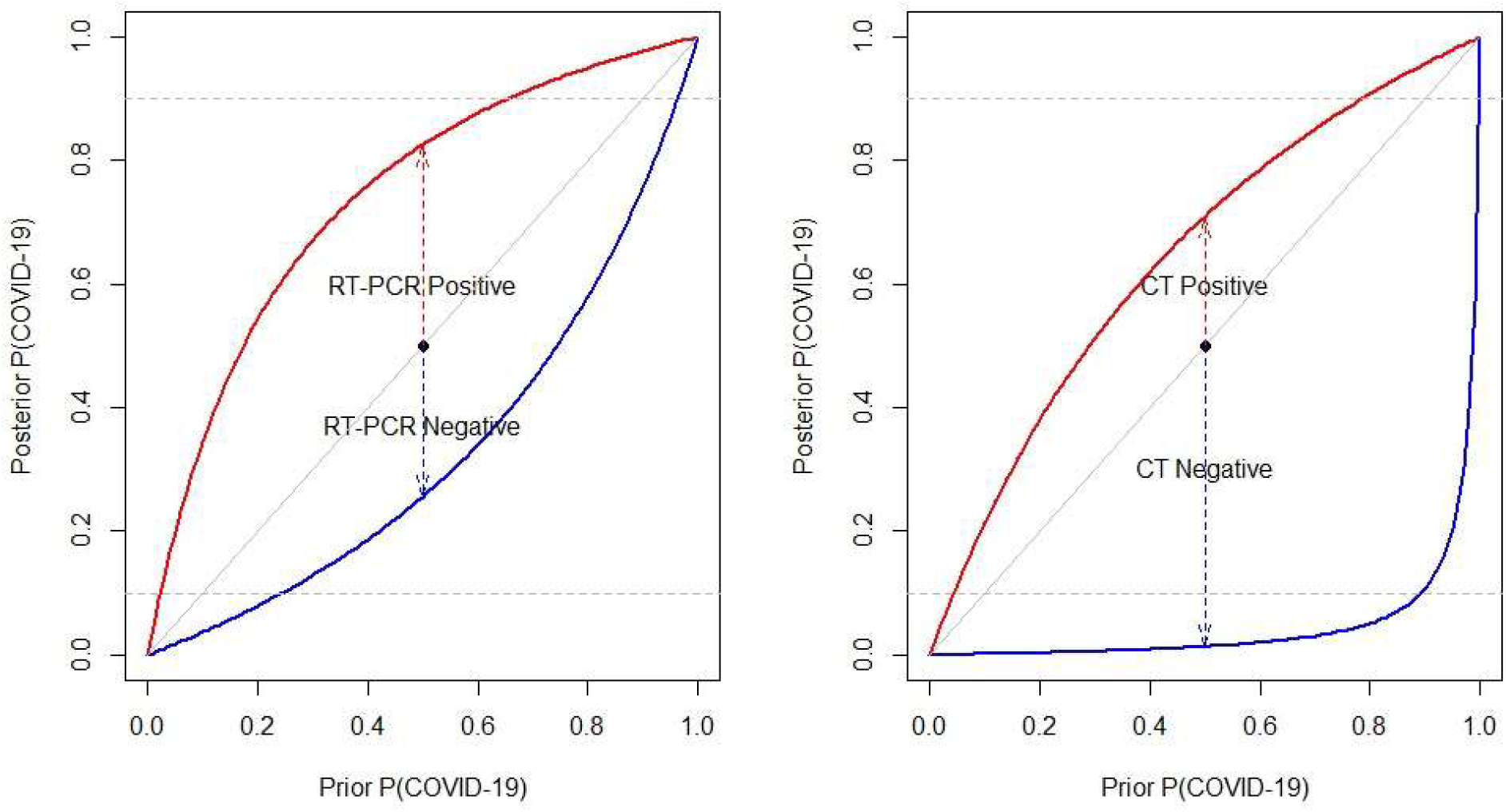
Probability of having COVID-19 predicted by RT-PCR (left) and chest CT (right) diagnostic tests as a function of the prior (pre-test) probability. The upper (red) curve applies for a person who tests positive, while the lower (blue) curve applies when the test is negative. The diagonal is the line of equality of pre-test and post-test probabilities; any point on it may be considered the predicted probability for a person before the diagnostic test is done. The arrows show an example of the changes induced by test results. For RT-PCR, the pre-test probability of 0.50 climbs to the post-test value of 0.83 after a positive test or it drops to 0.26 after a negative test result. For chest CT, the corresponding post-test probabilities of infection are 0.71 for a positive test or 0.01 for a negative test. The horizontal dashed lines mark probabilities of 10% and 90% for reference.

## Discussion

RT-PCR tests are commonly used for the diagnosis of many influenza viruses and coronaviruses. However, they are often treated as the gold standard in comparisons made to other diagnostic methods, which has led to a rarity of estimates of their diagnostic accuracy in clinical practice. The virus culture process is considered a better standard, but it takes several days instead of the few hours needed for RT-PCR tests. In one such comparison [16], RT-PCR was found to have sensitivity greater than 96% relative to virus culture for the diagnosis of H1N1 influenza. Similarly, high accuracy of RT-PCR has also been reported for the MERS coronavirus [10]. On the other hand, low accuracy has been reported for detection of SARS coronavirus with real-time RT-PCR [14, 20], although rates of detection were improved with the refinement of laboratory methods [22].

In the current COVID-19 pandemic, it has been a great boon to have had the rapid development of RT-PCR diagnostic tests that target the detection of different genes from the viral RNA. Laboratory testing has shown that at least one version of the RT-PCR assay can detect viral loads as small as 3.2 RNA copies per reaction [8] and that it does not cross-react to other known coronaviruses, particularly when the primer for the assay is well-chosen [6]. However, there is widespread doubt about how well the tests have worked in practice [12, 15]. Several sources of error were mentioned in the *Introduction* that include among them the uncertain distribution of the virus in the body at various times during the COVID-19 disease trajectory [27]. Comparisons of specimens from nasal and throat swabs indicate better sensitivity in nasal swabs and diminished sensitivity in throat swabs, particularly after the first few days of disease onset [31]. The variation in the severity of the viral infection between subjects presents another source of error with milder infections being more likely to escape detection.

The error rates of diagnostic testing using RT-PCR for COVID-19 that were estimated in this study may be considered to provide the cumulative impact of various sources of error. Our analysis showed 70.7% sensitivity of the RT-PCR diagnostic test that was used in Wuhan during the early stage of the pandemic, designed by China CDC and implemented with TaqMan One-Step RT-PCR kits. Even at the upper end of the 95% credible interval the sensitivity reached only 74.9%, which implies that the false negative rate exceeded 25%. The specificity of the test was better, estimated to be in the range from 77.4% to 94.1%. Using a reverse calculation that considered the chest CT as gold standard [11], the sensitivity and specificity of the same version of the RT-PCR test was reported to be 65% and 83%, respectively. Two other reverse calculations carried out by the same authors [11] on studies from China and Italy [5, 7] provided estimates as low as 47% for sensitivity and as high as 100% for specificity. However, the authors [11] acknowledged that the reverse calculations underestimated sensitivity and overestimated specificity of the RT-PCR tests, which is an expectation that is consistent with our findings.

Among reasons for the low sensitivity of RT-PCR might be that the Wuhan study used throat swabs rather than nasal swabs and that the optimal timing for testing was still in the process of being discovered. For example, a later study of disease propagation among 4950 quarantined Chinese participants reported that the first and second RT-PCR tests on throat-swab samples collected two days apart were positive in 72% and 92% of the 129 people who were eventually diagnosed with COVID-19 [17]. If data about the severity of infections and measures of viral load, such as cycle threshold of RT-PCR assays, had been available it might have been possible to explore whether it was the milder cases that tended to be misdiagnosed. It is likely that some of the diagnostic errors that resulted from the low sensitivity of RT-PCR were mitigated by the actions of medical professionals in Wuhan who might have decided to ignore negative test results in symptomatic patients. Nevertheless, the false negative rate is still likely to be among the main reasons for the difficulty in controlling the breakout in its early stages. For instance, if the prevalence is 10% in a population, our results indicate that testing would miss approximately 293 cases of COVID-19 for every 10,000 people tested. A highly transmissible virus can continue to propagate through the misdiagnosed cases.

Despite the focus of this investigation on the early version of the RT-PCR test that was used in Wuhan in January and February 2020, the estimated values of sensitivity and specificity have some resemblance to findings about other RT-PCR tests. An analysis of data from seven longitudinal studies found that the probability of false negatives of RT-PCR tests reached their lowest value of 20% on the third day after onset of symptoms before starting to increase again [13]. An Italian study of multiple RT-PCR tests that targeted different genes of viral RNA used a repeated testing design in the emergency room and reported sensitivity values ranging from 62% to 94% [4]. It was not clear whether they studied a test that may be considered equivalent to the Chinese test, but the range of reported sensitivity values includes the point estimate and error interval that was estimated in this study. Woloshin et al. [29] concluded that after consideration of current evidence, sensitivity and specificity values of 70% and 95% were reasonable estimates for RT-PCR tests. Although their estimate of specificity is high, the sensitivity value matches closely with our estimate. However, we also note that a systematic review found a high level of heterogeneity in the data for false negative rates [2], which serves as a warning against indiscriminate generalization of our results to other versions of RT-PCR tests.

Measurement of diagnostic accuracy of chest CT exams was not an aim of this study. However, the availability of data from chest CT exams alongside RT-PCR test results assisted in improving the estimation of diagnostic performance of the RT-PCR test. Moreover, our estimates for the high sensitivity (97.1% to 100%) and low specificity (48.2% to 70.8%) of chest CT were comparable to other reports. For example, using RT-PCR as gold standard, a study in Italy by Caruso et al. [5] reported 97% sensitivity and 56% specificity. On the other hand, Ai et al. [1] reported 96.5% sensitivity and 25.4% specificity in their study in Wuhan which was also based on assuming that RT-PCR was the gold standard test. We found a substantially higher estimate of specificity based on the same data, which can be attributed to the advantage of taking expert opinion into account and avoiding the comparison to a flawed gold standard test.

As far as the medical practitioner is concerned, the predictive values of the diagnostic test are of utmost importance. For COVID-19, if a medical practitioner suspected that there was a 50% pre-test chance that a patient had the disease, a subsequent negative RT-PCR test would mean that the patient still has 26% chance of the disease. A second confirmatory negative test would bring the chance of disease down to 10.5%, which continues to be an uncomfortably high chance. This illustrates some of the hardship of decision-making that was faced in Wuhan by the medical community.

From the methodological perspective, the approach adopted in this study delivered estimates of sensitivity and specificity that are free from the bias found in calculations based on an inaccurate gold standard test. The common reliance on a gold standard test was replaced here with the estimation of the prior distribution of the infected proportion coupled with Dawid and Skene’s [9] rigorous accounting of all possible true states of each patient. The utilization of expert opinion data in the form of verbal probabilities was key to the estimation of the prior proportion of COVID-19. Defuzzification made it possible to convert verbal probabilities to a framework that resides entirely within the domain of probability theory. The Bayesian computation flowed naturally from there on, with only a minor hurdle to overcome in marginalizing over the discrete set of true diagnoses to get past a limitation of the Hamiltonian sampling in *RStan*.

Among the many limitations of this study, the primary one is that the estimated sensitivity and specificity apply to the particular version of the RT-PCR test that was urgently created by China CDC [34] and that was being used in Wuhan, China, during January and February, 2020. An important methodological limitation of the study is that it is a retrospective study based on probabilistic knowledge of diagnostic errors and there were no studies available that were based on using a better gold standard test than RT-PCR that could act as a reference for comparison.

## Data Availability

Published data were used in this study.

## Acknowledgements

A previous version of this manuscript is available as a preprint [18].

## References

1. Ai T, Yang Z, Hou H, Zhan C, Chen C, Lv W, Tao Q, Sun Z, Xia L (2020) Correlation of Chest CT and RT-PCR Testing in Coronavirus Disease 2019 (COVID-19) in China: A Report of 1014 Cases. Radiology 200642. doi: 10.1148/radiol.2020200642

2. Arevalo-Rodriguez I, Buitrago-Garcia D, Simancas-Racines D, Zambrano-Achig P, Campo RD, Ciapponi A, Sued O, Martinez-García L, Rutjes AW, Low N, Bossuyt PM, Perez-Molina JA, Zamora J (2020) False-negative results of initial RT-PCR assays for COVID-19: A systematic review. PLOS ONE 15:e0242958. doi: 10.1371/journal.pone.0242958

3. Bélisle P, Joseph L (2017) beta.parms.from.quantiles, an R program for computing beta distribution parameters

4. Bisoffi Z, Pomari E, Deiana M, Piubelli C, Ronzoni N, Beltrame A, Bertoli G, Riccardi N, Perandin F, Formenti F, Gobbi F, Buonfrate D, Silva R (2020) Sensitivity, Specificity and Predictive Values of Molecular and Serological Tests for COVID-19: A Longitudinal Study in Emergency Room. Diagnostics 10:669. doi: 10.3390/diagnostics10090669

5. Caruso D, Zerunian M, Polici M, Pucciarelli F, Polidori T, Rucci C, Guido G, Bracci B, de Dominicis C, Laghi A (2020) Chest CT Features of COVID-19 in Rome, Italy. Radiology. doi: 10.1148/radiol.2020201237

6. Chan JF-W, Yip CC-Y, To KK-W, Tang TH-C, Wong SC-Y, Leung K-H, Fung AY-F, Ng AC-K, Zou Z, Tsoi H-W, Choi GK-Y, Tam AR, Cheng VC-C, Chan K-H, Tsang OT-Y, Yuen K-Y (2020) Improved molecular diagnosis of COVID-19 by the novel, highly sensitive and specific COVID-19-RdRp/Hel real-time reverse transcription-polymerase chain reaction assay validated in vitro and with clinical specimens. J Clin Microbiol. doi: 10.1128/JCM.00310-20

7. Cheng Z, Lu Y, Cao Q, Qin L, Pan Z, Yan F, Yang W (2020) Clinical Features and Chest CT Manifestations of Coronavirus Disease 2019 (COVID-19) in a Single-Center Study in Shanghai, China. AJR Am J Roentgenol 215:121–126. doi: 10.2214/AJR.20.22959

8. Corman VM, Landt O, Kaiser M, Molenkamp R, Meijer A, Chu DK, Bleicker T, Brünink S, Schneider J, Schmidt ML, Mulders DG, Haagmans BL, Veer B van der, Brink S van den, Wijsman L, Goderski G, Romette J-L, Ellis J, Zambon M, Peiris M, Goossens H, Reusken C, Koopmans MP, Drosten C (2020) Detection of 2019 novel coronavirus (2019-nCoV) by real-time RT-PCR. Eurosurveillance 25:2000045. doi: 10.2807/1560-7917.ES.2020.25.3.2000045

9. Dawid AP, Skene AM (1979) Maximum Likelihood Estimation of Observer Error-Rates Using the EM Algorithm. J R Stat Soc Ser C Appl Stat 28:20–28

10. Huh HJ, Kim JY, Kwon HJ, Yun SA, Lee MK, Ki CS, Lee NY, Kim JW (2017) Performance Evaluation of the PowerChek MERS (upE & ORF1a) Real-Time PCR Kit for the Detection of Middle East Respiratory Syndrome Coronavirus RNA. Ann Lab Med 37:494–498. doi: 10.3343/alm.2017.37.6.494

11. Kovács A, Palásti P, Veréb D, Bozsik B, Palkó A, Kincses ZT (2020) The sensitivity and specificity of chest CT in the diagnosis of COVID-19. Eur Radiol 1–6. doi: 10.1007/s00330-020-07347-x

12. Krumholz HM (2020) If You Have Coronavirus Symptoms, Assume You Have the Illness, Even if You Test Negative. N. Y. Times

13. Kucirka LM, Lauer SA, Laeyendecker O, Boon D, Lessler J (2020) Variation in False-Negative Rate of Reverse Transcriptase Polymerase Chain Reaction–Based SARS-CoV-2 Tests by Time Since Exposure. Ann Intern Med. doi: 10.7326/M20-1495

14. Lau LT, Fung Y-WW, Wong FP-F, Lin SS-W, Wang CR, Li HL, Dillon N, Collins RA, Tam JS-L, Chan PKS, Wang CG, Yu AC-H (2003) A real-time PCR for SARS-coronavirus incorporating target gene pre-amplification. Biochem Biophys Res Commun 312:1290– 1296. doi: 10.1016/j.bbrc.2003.11.064

15. Lazar K, Ryan A (2020) How accurate are coronavirus tests? Doctors raise concern about ‘false-negative’ results. BostonGlobe.com

16. López Roa P, Catalán P, Giannella M, García de Viedma D, Sandonis V, Bouza E (2011) Comparison of real-time RT-PCR, shell vial culture, and conventional cell culture for the detection of the pandemic influenza A (H1N1) in hospitalized patients. Diagn Microbiol Infect Dis 69:428–431. doi: 10.1016/j.diagmicrobio.2010.11.007

17. Luo L, Liu D, Liao X, Wu X, Jing Q, Zheng J, Liu F, Yang S, Bi B, Li Z, Liu J, Song W, Zhu W, Wang Z, Zhang X, Chen P, Liu H, Cheng X, Cai M, Huang Q, Yang P, Yang X, Han Z, Tang J, Ma Y, Mao C (2020) Modes of contact and risk of transmission in COVID-19 among close contacts. medRxiv 2020.03.24.20042606. doi: 10.1101/2020.03.24.20042606

18. Padhye NS (2020) Reconstructed diagnostic sensitivity and specificity of the RT-PCR test for COVID-19. medRxiv 2020.04.24.20078949. doi: 10.1101/2020.04.24.20078949

19. Pang J, Wang MX, Ang IYH, Tan SHX, Lewis RF, Chen JI-P, Gutierrez RA, Gwee SXW, Chua PEY, Yang Q, Ng XY, Yap RK, Tan HY, Teo YY, Tan CC, Cook AR, Yap JC-H, Hsu LY (2020) Potential Rapid Diagnostics, Vaccine and Therapeutics for 2019 Novel Coronavirus (2019-nCoV): A Systematic Review. J Clin Med 9. doi: 10.3390/jcm9030623

20. Peiris J, Chu C, Cheng V, Chan K, Hung I, Poon L, Law K, Tang B, Hon T, Chan C, Chan K, Ng J, Zheng B, Ng W, Lai R, Guan Y, Yuen K (2003) Clinical progression and viral load in a community outbreak of coronavirus-associated SARS pneumonia: a prospective study. The Lancet 361:1767–1772. doi: 10.1016/S0140-6736(03)13412-5

21. Pfefferle S, Reucher S, Nörz D, Lütgehetmann M (2020) Evaluation of a quantitative RT-PCR assay for the detection of the emerging coronavirus SARS-CoV-2 using a high throughput system. Eurosurveillance 25:2000152. doi: 10.2807/1560-7917.ES.2020.25.9.2000152

22. Poon LLM, Chan KH, Wong OK, Yam WC, Yuen KY, Guan Y, Lo YMD, Peiris JSM (2003) Early diagnosis of SARS Coronavirus infection by real time RT-PCR. J Clin Virol 28:233–238. doi: 10.1016/j.jcv.2003.08.004

23. R Core Team (2020) R: A Language and Environment for Statistical Computing. R Foundation for Statistical Computing, Vienna, Austria

24. CBEM Reusken, Broberg EK, Haagmans B, Meijer A, Corman VM, Papa A, Charrel R, Drosten C, Koopmans M, Leitmeyer K, ERLI-Net on behalf of E-L and (2020) Laboratory readiness and response for novel coronavirus (2019-nCoV) in expert laboratories in 30 EU/EEA countries, January 2020. Eurosurveillance 25:2000082. doi: 10.2807/1560-7917.ES.2020.25.6.2000082

25. RStudio Team (2019) RStudio: Integrated Development for R. RStudio, Inc., Boston, MA

26. Stan Development Team (2020) RStan: the R interface to Stan

27. Wang W, Xu Y, Gao R, Lu R, Han K, Wu G, Tan W (2020) Detection of SARS-CoV-2 in Different Types of Clinical Specimens. JAMA. doi: 10.1001/jama.2020.3786

28. Wintle BC, Fraser H, Wills BC, Nicholson AE, Fidler F (2019) Verbal probabilities: Very likely to be somewhat more confusing than numbers. PLoS ONE 14. doi: 10.1371/journal.pone.0213522

29. Woloshin S, Patel N, Kesselheim AS (2020) False Negative Tests for SARS-CoV-2 Infection — Challenges and Implications. N Engl J Med 383:e38. doi: 10.1056/NEJMp2015897

30. Wu F, Zhao S, Yu B, Chen Y-M, Wang W, Song Z-G, Hu Y, Tao Z-W, Tian J-H, Pei Y-Y, Yuan M-L, Zhang Y-L, Dai F-H, Liu Y, Wang Q-M, Zheng J-J, Xu L, Holmes EC, Zhang Y-Z (2020) A new coronavirus associated with human respiratory disease in China. Nature 579:265–269. doi: 10.1038/s41586-020-2008-3

31. Yang Y, Yang M, Shen C, Wang F, Yuan J, Li J, Zhang M, Wang Z, Xing L, Wei J, Peng L, Wong G, Zheng H, Liao M, Feng K, Li J, Yang Q, Zhao J, Zhang Z, Liu L, Liu Y (2020) Evaluating the accuracy of different respiratory specimens in the laboratory diagnosis and monitoring the viral shedding of 2019-nCoV infections. medRxiv 2020.02.11.20021493. doi: 10.1101/2020.02.11.20021493

32. Zadeh LA (1975) The concept of a linguistic variable and its application to approximate reasoning—I. Inf Sci 8:199–249. doi: 10.1016/0020-0255(75)90036-5

33. WHO Coronavirus. https://www.who.int/emergencies/diseases/novel-coronavirus-2019. Accessed 16 Apr 2020

34. Chinese National Institute for Viral Disease Control and Prevention. In: Primer Probes Detect. Nov. Coronavirus RT-PCR. http://ivdc.chinacdc.cn/kyjz/202001/t20200121_211337.html. Accessed 23 Apr 2020

35. WHO COVID-19 Technical Guidance. https://www.who.int/emergencies/diseases/novel-coronavirus-2019/technical-guidance/laboratory-guidance. Accessed 23 Apr 2020

